# Real-life performance of a COVID-19 rapid antigen detection test targeting the SARS-CoV-2 nucleoprotein for diagnosis of COVID-19 due to the Omicron variant

**DOI:** 10.1101/2022.02.02.22270295

**Authors:** Paula de Michelena, Ignacio Torres, Ángela Ramos-García, Victoria Gosalbes, Nidia Ruiz, Ana Sanmartín, Pilar Botija, Sandrine Poujois, Dixie Huntley, Eliseo Albert, David Navarro

**Affiliations:** Microbiology Service, Clinic University Hospital, INCLIVA Health Research Institute, Valencia, Spain; Health Care center Benimaclet, Health Department Clínico-Malvarrosa, Valencia, Spain; Health Care center Salvador Pau, Health Department Clínico-Malvarrosa, Valencia, Spain; Health Care center Serrería, Health Department Clínico-Malvarrosa, Valencia, Spain; Primary Health Directorate, Health Department Clínico-Malvarrosa, Valencia, Spain; Department of Microbiology, School of Medicine, University of Valencia, Valencia, Spain

**Keywords:** SARS-CoV-2, Omicron variant, rapid antigen assay, COVID-19, field performance study

## Abstract

**Objectives:** It has been suggested that rapid antigen detection assays (RADT) may perform suboptimally in terms of sensitivity for the diagnosis of SARS-CoV-2 Omicron variant infection. To address this issue, we conducted a prospective study in primary health centers to evaluate the clinical performance of the Panbio™ COVID-19 Ag Rapid Test Device in nasopharyngeal specimens (NP) carried out at the point of care.

**Methods:** We recruited 244 patients (median age, 40 years; range 2–96; 141 female) with clinical suspicion of COVID-19 (232 adults and 12 children). 228/244 patients had been fully vaccinated (two doses) with licensed COVID-19 vaccines prior to recruitment. Most patients (222/244) were SARS-CoV-2 naïve prior to enrollment. Patients were tested by RT-PCR and RADT within 5 days since symptoms onset.

**Results:** 126 patients (51.6%) tested positive by both RT-PCR and RADT, 90 patients (36.8%) returned negative results by both assays and 28 patients (11.4%) yielded discordant results (RT-PCR+/RADT-). No patients tested RT-PCR-/RADT+. Overall specificity and sensitivity of RADT was 100% (95% CI, 95.9–100%) and 81.8% (95% CI, 75–87.1%) respectively. The sensitivity of the assay increased from 79.6% (95% CI, 66.4–88.5) when considering specimens collected at days 0–1 after symptoms onset, to 86.4% (95% CI, 66.7–95.3) when grouping the specimens obtained on days 4–5.

**Conclusion:** The Panbio™ COVID-19 Ag Rapid Test Device perform well (≥80% sensitivity) as a point-of-care test for early diagnosis of COVID-19 due to the Omicron variant in primary healthcare centers.

## INTRODUCTION

A number of commercially available rapid antigen detection tests (RADT) targeting the SARS-CoV-2 nucleocapsid protein (NP) have been shown to display a sensitivity of over 80% compared to RT-PCR assays for diagnosis of SARS-CoV-2 infection in symptomatic individuals, provided that testing is conducted within one week after symptoms onset [1]. These RADT were optimized for detection of the ancestral Wuhan-Hu-1 variant, and the emergence of SARS-CoV-2 variants which incorporate non-synonymous mutations within the amino acid sequence of NP may impact on the diagnostic efficiency of RADT. In this context, two studies [2,3] showed that the Panbio™ COVID-19 Ag Rapid Test Device (Abbott Diagnostic GmbH, Jena, Germany) had decreased sensitivity for detection of SARS-CoV-2 Alpha (B.1.1.7) variant compared to non-alpha lineages. As in many other countries, the SARS-CoV-2 Omicron variant has overtaken the Delta variant and currently dominates in Spain. It has been suggested that RADT may be less sensitive for detecting the Omicron variant [4], but this assumption lacks support by real-life RADT performance studies. Here, we conducted a prospective study in primary health centers to evaluate the clinical performance of the Panbio™ COVID-19 Ag Rapid Test Device in nasopharyngeal specimens (NP) carried out at the point of care for diagnosis of Omicron variant COVID-19.

## METHODS

### Patients

In the current observational prospective study, a convenience sample of 244 patients (median age, 40 years; range 2–96; 141 female) with clinical suspicion of COVID-19, 232 of whom were adults (median age, 41 years; range, 17–96) and 12 children (median age, 15 years; range, 2–16), attending three randomly selected primary care centers affiliated to the Clínico-Malvarrosa Health Department in Valencia (Spain) were recruited between January 10 and January 21. Only patients with symptoms developing within the previous 5 days were enrolled. The study was approved by the Hospital Clínico de Valencia (HCU) INCLIVA Research Ethics Committee. Since the testing strategy was considered as regular clinical practice according to local health authorities, written informed consent was waived by this committee.

### SARS-CoV-2 testing

We collected two NPs per patient, one of which (provided by the manufacturer) was used for RADT while the other was placed in 3□mL of universal transport medium (DeltaSwab Virus, Deltalab, Barcelona, Spain) and delivered to the HCU Microbiology Service for RT-PCR testing. RADT was performed immediately after sampling following the manufacturer’s instructions (reading at 15 min). RT-PCRs were carried out within 24 h of specimen collection with the TaqPath COVID-19 Combo Kit (Thermo Fisher Scientific, MS, USA) which targets SARS-CoV-2 ORF1ab, N and S genes. RNA was extracted using the Applied Biosystems™ MagMAX™ Viral/Pathogen II Nucleic Acid Isolation Kits coupled with Thermo Scientific™ KingFisher Flex automated instrument. The AMPLIRUN® TOTAL SARS-CoV-2 Control (Vircell SA, Granada, Spain) was used as the reference material for SARS-CoV-2 RNA load quantification [5], for which purpose RT-PCR cycle threshold (C_T_) values returned by amplification of the NP gene were considered. The S-gene dropout RT-PCR profile was systematically associated with the Omicron variant within the study period, as confirmed by whole-genome sequencing (not shown).

### Statistical analyses

Agreement between RAD assay and RT-PCR was assessed using Cohen’s κ statistics. Differences between medians were compared using the Mann–Whitney U-test. Two-sided *P* values < 0.05 were considered significant. Statistical analyses were performed using SPSS version 25.0 (SPSS, Chicago, IL, USA).

## RESULTS

Results relevant to interpretation of the data presented herein are as follows. First, 228 of the 244 patients had been fully vaccinated (two doses) with licensed COVID-19 vaccines prior to recruitment. Second, most patients (222 out of 244) were SARS-CoV-2 naïve prior to enrollment. In all, 126 patients (51.6%) tested positive by both RT-PCR and RADT and 90 patients (36.8%) returned negative results by both assays. In turn, 28 patients (11.4%) yielded discordant results (RT-PCR+/RADT-). No patients tested RT-PCR-/RADT+. Concordance between the results provided by the two assays was good (κ, 0.78; 95% CI, 0.69–0.85). Importantly, time to specimen collection was comparable (*P*=0.69) between RT-PCR+/RADT+ and RT-PCR+/RADT-patients (median 2 days; range, 0–5).

Overall specificity and sensitivity of RADT was 100% (95% CI, 95.9–100%) and 81.8% (95% CI, 75–87.1%) respectively. As shown in Table 1, RADT assay sensitivity increased in parallel with SARS-CoV-2 RNA load, reaching 95.6% in specimens with viral loads ≥7.5 log10 copies/ml (C_T_, ≤20). Interestingly, the sensitivity of the assay increased from 79.6% (95% CI, 66.4–88.5) when considering specimens collected at days 0–1 after symptoms onset, to 86.4% (95% CI, 66.7–95.3) when grouping the specimens obtained on days 4–5 (Supplementary Table 1).

**Table 1.**
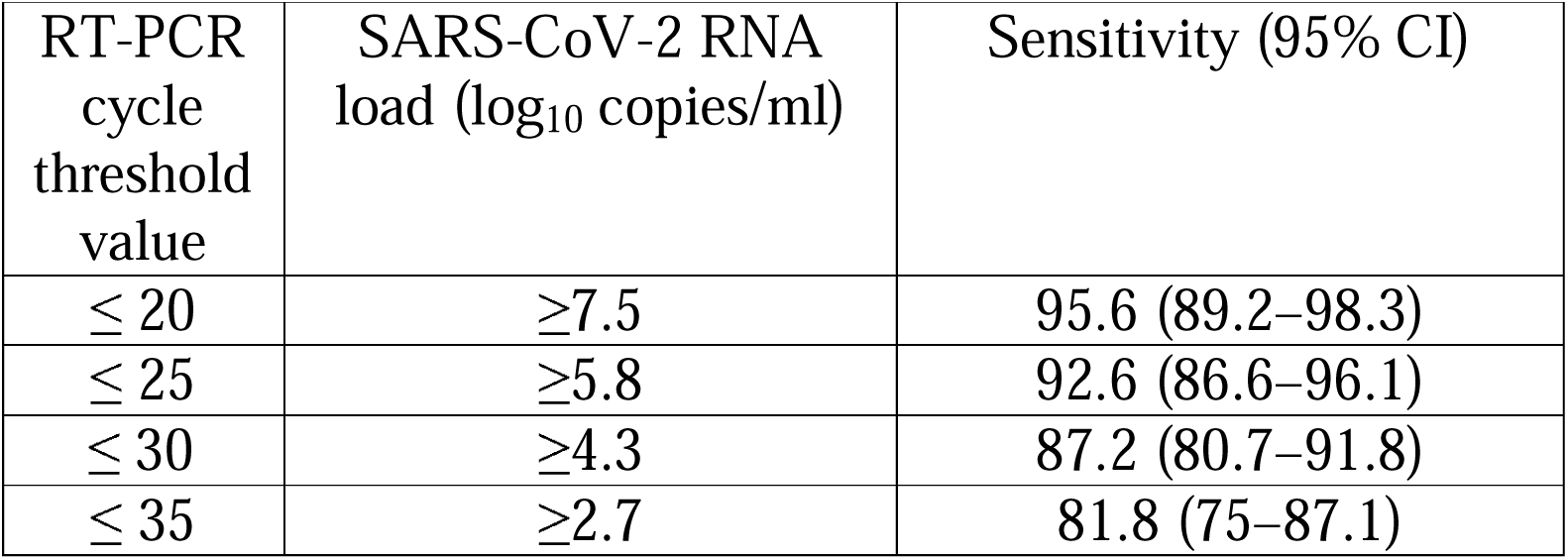
Overall sensitivity of the Panbio™ COVID-19 Ag Rapid Test Device according to the SARS-CoV-2 RNA load in nasopharyngeal specimens.

|  |  |  |
| --- | --- | --- |
| RT-PCR cycle threshold value | SARS-CoV-2 RNA load (log<sub>10</sub> copies/ml) | Sensitivity (95% CI) |
| ≤ 20 | ≥7.5 | 95.6 (89.2–98.3) |
| ≤ 25 | ≥5.8 | 92.6 (86.6–96.1) |
| ≤ 30 | ≥4.3 | 87.2 (80.7–91.8) |
| ≤ 35 | ≥2.7 | 81.8 (75–87.1) |

Overall, RADT negative predictive value for an estimated prevalence of 30% and 35% (representative of our Health Department during the study period) was 92.8% (95% CI, 85.8–96.5) and 91.1% (95% CI, 82.8–95.6), respectively.

As expected, median viral RNA load was significantly higher (*P*<0.0001) in RT-PCR+/RADT+ specimens than in those returning RT-PCR+/RADT-results (Fig. 1).

**Figure 1.**
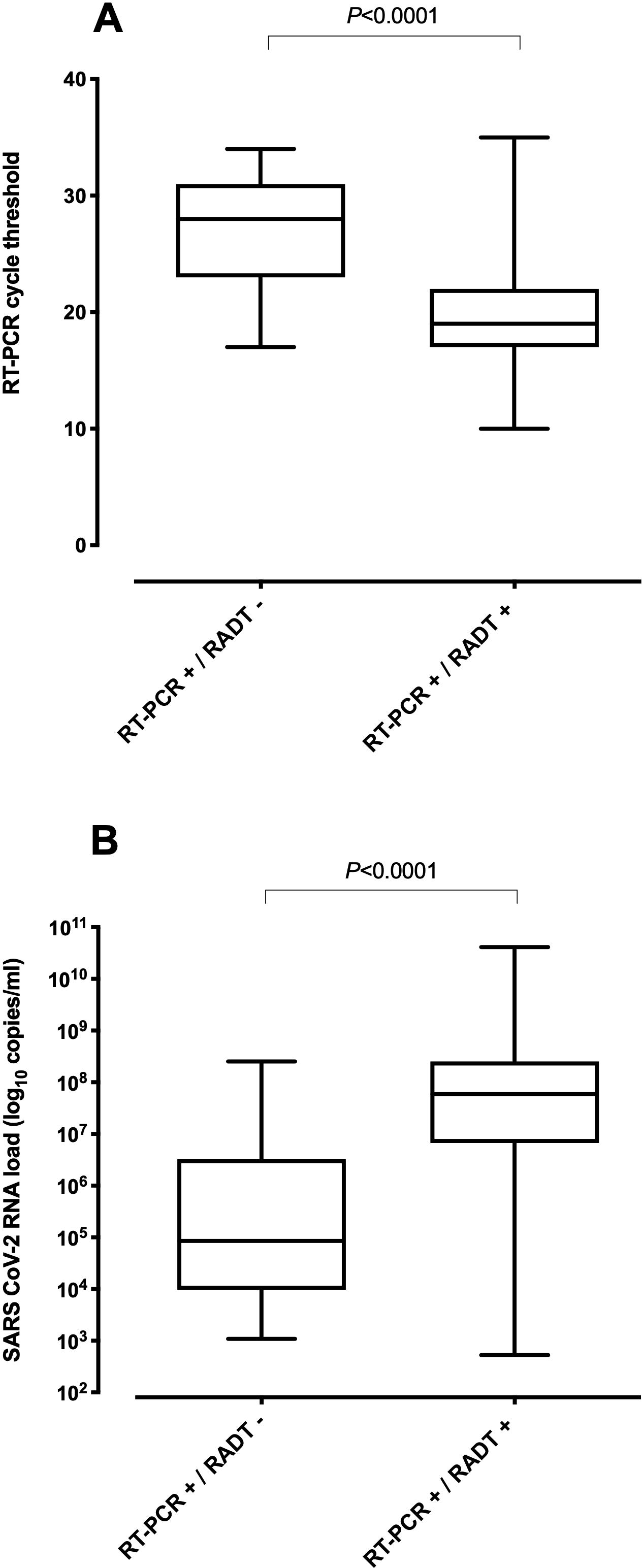
Box-Whisker plots depicting RT-PCR cycle threshold values (C_T_) (A) and viral RNA loads (B) in nasopharyngeal specimens collected from COVID-19 patients infected with the Omicron variant testing either positive or negative by the Panbio™ COVID-19 Ag Rapid Test Device (Abbott Diagnostic GmbH, Jena, Germany). *P* values for comparisons are shown.

## DISCUSSION

The SARS-CoV-2 Omicron variant carries one or more mutations in the NP gene (P13L, Del31-33, R203K and G203K) [6] that may impact on the sensitivity of RADT [4]. The Abbott BinaxNow test (similar to the Panbio™ COVID-19 assay) performed on nasal specimens was reported to produce false negative results in four individuals who nonetheless were confirmed to have transmitted the Omicron variant to close contacts [7]. Likewise, the Panbio™ COVID-19 assay was found to perform worse for diagnosis of COVID-19 due to vaccine-breakthrough Omicron infection (sensitivity of 36.1%) compared to Delta (sensitivity of 67.7%) [8]. Nevertheless, in that study [8], NP were diluted in viral transport medium and cryopreserved prior to RADT testing. When using live virus isolated from clinical specimens, the Panbio™ COVID-19 assay displayed comparable analytical sensitivity for detection of Omicron and Delta in one study [9], but lower for Omicron in another [8]. To our knowledge, no published studies have evaluated the performance of RADT conducted at point of care for diagnosis of the Omicron variant of COVID-19. In a series comprising mostly vaccinated adult individuals with no history of SARS-CoV-2 infection prior to enrollment and tested within 5 days after symptoms onset, we showed the Panbio™ COVID-19 Ag Rapid Test Device to display exquisite specificity and acceptable overall sensitivity (81.8%) for Omicron diagnosis, both figures exceeding regulatory agency requirements for temporary approval (at least 98% and 80%, respectively) [1]. In our experience, the clinical performance of the Panbio™ COVID-19 assay for Omicron variant was comparable to previous reports for the Wuhan-Hu-1 G614 variant (100% specificity and sensitivity of 81.4% in non-vaccinated adult patients with a clinical course of <5 days) [5]. In line with previous findings from our group [5], RADT sensitivity increased in parallel with SARS-CoV-2 RNA load in NP. Interestingly, the sensitivity of the RADT assay also appeared to increase with time elapsed after symptoms onset, suggesting that vaccine-breakthrough Omicron variant infection may be symptomatic even in the presence of RNA loads below the threshold for viral detection by RADT. Although speculative, this phenomenon may be related to the reduced capability of the Omicron variant to antagonize the host cell interferon response [10].

The limitations of the current study are as follows. First, Omicron subvariant B.1.529.2 (BA.2), which lacks the 69-70 deletion, may be incorrectly categorized as such based on the SGTF result; nonetheless, this subvariant could not be identified in sequenced specimens within the study period in our health department. Second, side-by-side clinical performance comparison of the RADT for diagnosis of COVID-19 due to Omicron and other variants of concern was not possible due to the absolute dominance of the former at the time of study. Third, no cell cultures were performed for specimens returning discordant RT-PCR/RADT results. Fourth, the small number of children, SARS-CoV-2-experienced and unvaccinated individuals enrolled precluded conducting robust subanalyses for these population groups.

In summary, we found the Panbio™ COVID-19 Ag Rapid Test Device to perform well as a point-of-care test for early diagnosis of COVID-19 due to the Omicron variant in primary healthcare centers. Further studies are warranted to evaluate the performance of this and other RADT for detection of Omicron variant infection in asymptomatic, pediatric and unvaccinated individuals.

## Data Availability

The data that support the findings of this study are available on request from the corresponding author

## ACKNOWLEDGMENTS

We are grateful to residents and staff at the Microbiology Service of Hospital Clínico Universitario and medical and nursing staff at primary health centers (Benimaclet, Serrería and Salvador Pau) recruiting patients for the current study. Ignacio Torres (Río Hortega Contract; CM20/00090) and Eliseo Albert (Juan Rodés Contract; JR20/00011) hold contracts funded by the Carlos III Health Institute (co-financed by the European Regional Development Fund, ERDF/FEDER).

## AUTHOR CONTRIBUTIONS

PdM, IT, SP, DH, EA, methodology and data validation. AR-G, VG, NR, AS and PB, study design and logistics; DN, conceptualization, data analysis and manuscript writing.

## FIGURE LEGENDS

**Supplementary Table 1.** SARS-CoV-2 RT-PCR and Panbio™ COVID-19 Ag Rapid Test results in nasopharyngeal specimens collected from COVID-19 patients according to time elapsed since onset of symptoms.

| Panbio™ COVID-19 Ag Rapid Test result in study groups | SARS-CoV-2 RT-PCR result |  |
| --- | --- | --- |
|  | No. of positives | No. of negatives |
| **All patients (n=244)** |  |  |
| Positive | 126 | 0 |
| Negative | 28 | 90 |
| **Patients tested days 0-1 after symptoms onset (n=71)** |  |  |
| Positive (n=39) | 39 | 0 |
| Negative (n=10) | 10 | 21 |
| **Patients tested days 2-3 after symptoms onset (n=129)** |  |  |
| Positive (n=68) | 68 | 0 |
| Negative (n=15) | 15 | 46 |
| **Patients tested days 4-5 after symptoms onset (n=44)** |  |  |
| Positive (n=19) | 19 | 0 |
| Negative (n=3) | 3 | 22 |

## REFERENCES

1. Drain PK. Rapid Diagnostic Testing for SARS-CoV-2. N Engl J Med 2022 Jan 7. doi: 10.1056/NEJMcp2117115.

2. Jian MJ, Chung HY, Chang CK, Lin JC, Yeh KM, Chen CW, et al. ARS-CoV-2 variants with T135I nucleocapsid mutations may affect antigen test performance. Int J Infect Dis 2022;114:112–114.

3. van Ogtrop ML, van de Laar TJW, Eggink D, Vanhommerig JW, van der Reijden WA Comparison of the Performance of the PanBio COVID-19 Antigen Test in SARS-CoV-2 B.1.1.7 (Alpha) Variants versus non-B.1.1.7 Variants. Microbiol Spectr 2021; 9:e0088421.

4. https://www.fda.gov/medical-devices/coronavirus-covid-19-and-medical-devices/sars-cov-2-viral-mutations-impact-covid-19-tests

5. Albert E, Torres I, Bueno F, Huntley D, Molla E, Fernández-Fuentes MÁ, et al. Field evaluation of a rapid antigen test (Panbio™ COVID-19 Ag Rapid Test Device) for COVID-19 diagnosis in primary healthcare centres. Clin Microbiol Infect 2021;27:472.e7-472.e10.

6. https://covdb.stanford.edu/page/mutation-viewer/#omicron.

7. Adamson B, Sikka R, Wyllie AL, Premsrirut P. Discordant SARS-CoV-2 PCR and Rapid Antigen Test Results When Infectious: A December 2021 Occupational Case Series. medRxiv 2022.01.04.22268770; doi: https://doi.org/10.1101/2022.01.04.22268770.

8. Bekliz M, Perez-Rodriguez F, Puhach O, Kenneth Adea, Marques Melancia S, Baggio S, et al. Sensitivity of SARS-CoV-2 antigen-detecting rapid tests for Omicron variant. medRxiv 2021.12.18.21268 018; doi: https://doi.org/10.1101/2021.12.18.21268018.

9. Deerain J, Druce J, Tran T, Batty M, Yoga Y, Fennell M, et al. Assessment of the analytical sensitivity of ten lateral flow devices against the SARS-CoV-2 omicron variant. J Clin Microbiol 2021;Dec 22:jcm0247921.

10. Bojkova D, Widera M, Ciesek S, Wass MN, Michaelis M, Cinatl jr. J. Reduced interferon antagonism but similar drug sensitivity in Omicron variant compared to Delta variant SARS-CoV-2 isolates. BioxRiv 2022; doi: https://doi.org/10.1101/2022.01.03.474773.

